# PhysiCase: Development and dual-layer validation of synthetic cases for health professional education: A pilot study leveraging Generative AI

**DOI:** 10.64898/2026.06.07.26355114

**Authors:** Oyindolapo O. Komolafe, Angela C. Roberts, Jacob Shelley, Andrews Tawiah

## Abstract

High-quality, domain-specific datasets are foundational to advancing educational tools and AI systems in healthcare, yet assembling case repositories from real-world clinical records faces substantial privacy, ethical, and licensing barriers. Synthetic data generation offers a compelling pathway forward, but educational cases require rigorous validation to ensure clinical plausibility and pedagogical utility. This pilot study introduces PhysiCase, a dual-layer validation pipeline for synthetic case generation and evaluates the feasibility of combining automated LLM-based screening with expert educator review.

We generated 128 synthetic musculoskeletal(MSK) cases using four frontier large language models (GPT-4.1, GPT-4o, Google Gemini 2.5 Pro, and Llama 4 Scout) across 28 clinical conditions. Cases underwent automated quality screening using an “LLM-as-judge” framework (DeepEval) assessing prompt alignment, JSON correctness, answer relevance, bias, toxicity, and completeness. Ninety cases (70.3%) passed automated filtering and proceeded to expert evaluation by four MSK physiotherapy educators, who rated medical accuracy, realism, fidelity, relevance, and usability on 5-point Likert scales.

GPT-4.1 demonstrated the highest automated pass rate (96%) and strongest expert ratings (medical accuracy 4.10/5, usability 4.38/5), while Llama 4 Scout showed the lowest pass rate (33.3%) and expert ratings. Expert-evaluated cases achieved strong content validity indices for usability (97.5%), relevance (97.5%), and realism (95%), though medical accuracy showed greater variance (CVI 87.5%). Cross-layer correlation analysis revealed that automated completeness metrics moderately aligned with expert usability ratings (*ρ* ≈ 0.393), while answer relevance and prompt alignment showed weak or negative correlations with clinical correctness. Qualitative analysis identified three primary failure modes: reductive logic, biomechanical inconsistency, and administrative/contextual gaps.

The dual-layer validation framework proved methodologically viable: automated screening efficiently reduced expert review burden, while human judgment remained indispensable for detecting subtle clinical reasoning failures. LLM-generated synthetic cases has the potential to meet practical educational needs for MSK physiotherapy, but expert validation is essential to safeguard clinical accuracy. These findings support a scalable division of labour for synthetic case development, with targeted improvements to prompting and automated reasoning checks needed to address identified “nuance gaps.”

The code for this paper is available on GitHub.

## 1 Introduction

High-quality, domain-specific datasets are foundational to advancing both educational tools and AI systems in healthcare [1, 2]. In health professional education specifically physiotherapy, effective training depends on exposure to diverse case material that captures authentic clinical reasoning, presentation variability, and context-sensitive decision-making [3, 4]. Yet assembling such case repositories from real-world clinical records faces substantial barriers including privacy regulations, institutional restrictions, and ethical constraints governing patient-derived data [5]. Compounding these challenges, many existing case banks operate under restrictive licensing that explicitly prohibits AI training or algorithmic generation, effectively blocking their use for modern data-centric educational applications. This dual constraint—limited access to real cases and restricted use of existing resources—creates a critical bottleneck for educators and researchers seeking large, diverse corpora for training both students and machine learning models.

Synthetic data generation offers a compelling pathway forward. By creating artificial patient cases that replicate authentic clinical scenarios without exposing health information, synthetic data can bridge access gaps in research and education [6–9]. However, the central challenge is not generation of case studies but their validation [6–9]. Unlike synthetic data for administrative or research purposes, educational cases must achieve high fidelity to clinical reality; low-quality synthetic cases risk active harm by encoding incorrect pathophysiological mechanisms, misleading imaging interpretations, or unrealistic pain presentations that teach learners fundamentally flawed clinical abstractions [10, 11]. Consequently, any synthetic dataset intended for educational reuse requires rigorous evaluation frameworks that combine scalable automated screening with domain-expert verification. Large language models (LLMs) have recently demonstrated substantial capability in generating human-like clinical text, and medicine has begun exploring their potential for training applications [12, 13].

This paper presents PhysiCase, a pilot study introducing and evaluating a dual-layer validation pipeline for case generation. We operationalize a three-phase workflow that: (i) generates structured cases using multiple frontier LLMs; (ii) applies automated filtering through an “LLM-as-judge” quality gate; and (iii) subjects retained cases to expert educator review with structured ratings and qualitative feedback. This pilot domain is physiotherapy musculoskeletal (MSK) cases and it hopes to approach an understanding of the following questions:

- **RQ1 (Educational validity)**: To what extent do automatically filtered synthetic cases exhibit the clinical plausibility and pedagogical utility required for professional education?
- **RQ2 (Metric alignment)**: How well do automated evaluation metrics (e.g., prompt alignment, completeness, relevance) correlate with expert assessments of medical accuracy and usability?

Our work makes three primary contributions: (1) the PhysiCase pilot dataset - synthetic MSK physiotherapy cases spanning multiple conditions and LLM generators, annotated with automated metrics and expert ratings; (2) a reproducible dual-layer validation pipeline that balances scalable automated screening with expert review as the criterion standard for clinical validity; and (3) empirical evidence on nuance gaps—specific expert-identified failure themes that directly inform subsequent development phases.

## 2 Related Work

### 2.1 Synthetic Data Use in Healthcare

In healthcare, the use of synthetic data is seen as a transformative solutions to critical challenges related to data privacy, scarcity, and accessibility, although its large-scale adoption is currently hampered by technical and regulatory barriers [9, 14].

Giuffrè and Shung [15] provide a comprehensive overview of synthetic data applications in healthcare, demonstrating how artificially generated datasets can enhance data privacy while maintaining utility for predictive analytics. Their work establishes that synthetic data serves three primary functions in healthcare: preserving patient privacy, augmenting limited datasets for machine learning, and enabling cross-institutional collaboration without exposing sensitive information. In medical imaging specifically, synthetic data has gained substantial traction. Koetzier et al. [16] conducted a systematic review examining AI-generated synthetic medical imaging data, finding that generative models can produce high-fidelity images that address both data scarcity and privacy concerns while maintaining diagnostic utility. The authors emphasize that synthetic data generation has been tested across multiple imaging modalities including radiography, computed tomography, magnetic resonance imaging, and ultrasound, with promising results for training diagnostic algorithms.

The privacy-preserving potential of synthetic data is particularly significant given stringent regulatory requirements. Liu, Acharya, and Tan [17] conducted a systematic review of deep learning approaches for synthetic data generation in healthcare, focusing specifically on privacy preservation capabilities. Their analysis reveals that while synthetic data reduces re-identification risks compared to anonymization techniques, challenges remain in ensuring that generated data does not leak information about individual patients in the training set. This concern is echoed by Arora et al. [18] who argue for accelerated development of synthetic data privacy frameworks to address person-level identification risks that persist even in synthetic medical images.

Recent advances in generative AI have expanded synthetic data capabilities substantially. Van Breugel et al. [19] review generative AI applications for biomedical synthetic data, covering tabular data, images, and graphs for diverse bioengineering and medical applications. Their analysis demonstrates that diffusion models and transformer-based architectures have significantly improved the fidelity and diversity of synthetic healthcare data. Similarly, Wang et al. [20] developed a self-improving generative foundation model for synthetic medical image generation, demonstrating that AI models trained on both real and synthetic data exhibit improved predictive capabilities, particularly benefiting rare disease detection where real data is scarce .

However, quality assurance remains a critical concern. Kaabachi et al. [21] conducted a scoping review of privacy and utility metrics in medical synthetic data, finding that while numerous metrics exist for assessing synthetic data utility, no consensus has emerged on standardized approaches to evaluate privacy protection. This gap in standardization complicates regulatory approval and clinical adoption of synthetic data solutions. Yale et al. [22] previously established evaluation frameworks for privacy-preserving synthetic health data using novel metrics to capture dataset resemblance and privacy preservation, providing foundational methodologies still referenced in current research.

The European Health Data Space (EHDS) has recognized synthetic data’s potential, with Jiang, Domingues, and Mendes [23] examining how synthetic data can address privacy preservation, data scarcity, and bias mitigation within cross-border healthcare data sharing frameworks . Their analysis presents use cases demonstrating synthetic data’s role in enabling research while maintaining compliance with evolving data protection regulations.

### 2.2 LLM Use in Physiotherapy context

Large language models (LLMs) are increasingly being explored for applications in physiotherapy and physical rehabilitation, though this domain remains relatively nascent compared to other medical specialties. Naqvi, Shaikh, and Mishra [24] provide a foundational perspective on LLM integration in physical therapy practice, arguing that these technologies present unique opportunities for physiotherapists to elevate clinical practice through enhanced documentation, patient education, and clinical decision support.

Hao et al. [25] evaluated ChatGPT’s role in clinical decision-making for MSK physical therapy, finding that GPT-4 can provide accurate and detailed suggestions that align with clinical practice guideline. Their study specifically examined ChatGPT’s performance in assisting diagnostic reasoning and treatment planning for MSK conditions, establishing baseline capabilities for AI-assisted physiotherapy practice.

Comparative evaluations reveal performance variations across LLM versions. Sağlam et al. [26] conducted a cross-sectional study comparing GPT-4 and GPT-3.5 in clinical decision-making for sports surgery and physiotherapy, demonstrating that GPT-4 significantly outperforms its predecessor in clinical reasoning tasks. This finding has important implications for physiotherapy education and practice, suggesting that newer model generations offer substantially improved clinical utility.

In education applications, Ferrer-Peña et al. [27] conducted a randomized controlled trial examining LLM-based clinical reasoning training for physical therapy students, representing the first study to implement LLM technology specifically for improving clinical reasoning skills in physiotherapy education.

Their pilot study establishes proof-of-concept for AI-augmented physiotherapy education, though they note the need for larger trials to validate educational outcomes. Almeida et al. [28] developed Physio, an LLM-based physiotherapy advisor capable of making initial diagnoses while citing reliable health sources . This chat-based application demonstrates how LLMs can be constrained to provide verifiable information through retrieval-augmented generation, combining generative language capabilities with external knowledge databases to recommend rehabilitation exercises and over-the-counter medications.

However, significant limitations persist, as shown in Safran and Yildirim’s [29] evaluation of ChatGPT’s alignment with clinical practice guidelines in MSK rehabilitation, finding that while GPT-4 demonstrates reasonable performance, gaps remain in addressing complex clinical scenarios requiring contextual judgment. Their study emphasizes that therapist-patient interaction and nuanced clinical reasoning remain areas where human expertise surpasses current LLM capabilities. Rosen et al. [30] explored physical therapists’ perspectives on LLM-powered knowledge translation tools for guideline adherence, finding that practitioners view these technologies as potentially valuable for improving evidence-based practice. Their qualitative focus group study reveals both enthusiasm for AI-assisted decision support and concerns regarding the quality and reliability of LLM-generated recommendations. Michou et al. [31] compared LLM performance against physiotherapy students in clinical rehabilitation scenarios within the Greek healthcare context, finding that while LLMs demonstrate knowledge gains, they overlook important elements of hands-on clinical training emphasized in European physiotherapy curricula. This research highlights the complementary rather than replacement role of AI in physiotherapy education.

### 2.3 The Use of LLM to Generate Synthetic Data

The intersection of LLMs and synthetic data generation represents a rapidly evolving research frontier with significant implications for healthcare AI development. Ding et al. [32] provided a comprehensive survey of data augmentation using LLMs, examining data perspectives, learning paradigms, and challenges associated with training models on LLM-generated data. Their analysis revealed that LLM-based data augmentation differs fundamentally from traditional techniques by generating semantically coherent synthetic examples rather than simply perturbing existing data points.

Whitehouse, Choudhury, and Aji [33] leveraged LLMs to generate synthetic data for enhanced cross-lingual performance, showing that LLM-generated synthetic examples can substantially improve model performance in low-resource languages. This approach has particular relevance for healthcare applications where multilingual clinical data is scarce. Kang et al. [34] developed an LLM-based approach for generating synthetic data to improve depression prediction, demonstrating that synthetic data augmentation can address data scarcity and privacy concerns while enhancing dataset utility. Their novel approach illustrates how domain-specific synthetic data generation can augment limited clinical datasets for mental health applications.

However, significant challenges persist regarding bias inheritance. Li et al. [35] examined bias propagation in LLM-based data augmentation, finding that when LLMs generate synthetic training data, they may amplify existing biases present in their training corpora. Their analysis demonstrates that bias ratios in augmented datasets require careful monitoring to prevent downstream task degradation, particularly critical in healthcare where biased synthetic data could perpetuate health disparities. He et al. [36] conducted a critical analysis of AI-generated data contamination, examining over 800,000 synthetic data points across clinical text generation, vision-language reporting, and medical image synthesis. Their alarming findings indicate that without mandatory human verification, iterative training on AI-generated data drives rapid erosion of pathological variability, with rare but critical findings vanishing from synthetic content and false reassurance rates tripling to 40%. This research underscores the critical importance of human oversight in synthetic data pipelines.

The convergence of LLMs and synthetic data generation presents both opportunities and risks for healthcare AI. While LLMs enable scalable generation of diverse, semantically coherent synthetic examples, He et al.’s [36] findings regarding model collapse and diagnostic reliability degradation serve as a critical caution against uncritical adoption . The research collectively suggests that LLM-generated synthetic data requires rigorous validation, quality-aware filtering, and maintained human oversight to realize its potential while avoiding degradation of healthcare data ecosystems. Therefore, this work presents a dual-layer validation pipeline for validating synthetic case-study data generated by LLMs for MSK physiotherapy education.

## 3 Method

### 3.1 PhysiCase Pipeline

NVIDIA’s generative synthetic data workflow [37] was adapted to the task of creating text-based clinical case studies. Our adapted pipeline (Figure 1) consists of several phases: 1. Data Generation, 2. Automated Filtering, and 3. Human Evaluation. Key components and decisions in this pipeline are detailed below.

**Fig 1.**
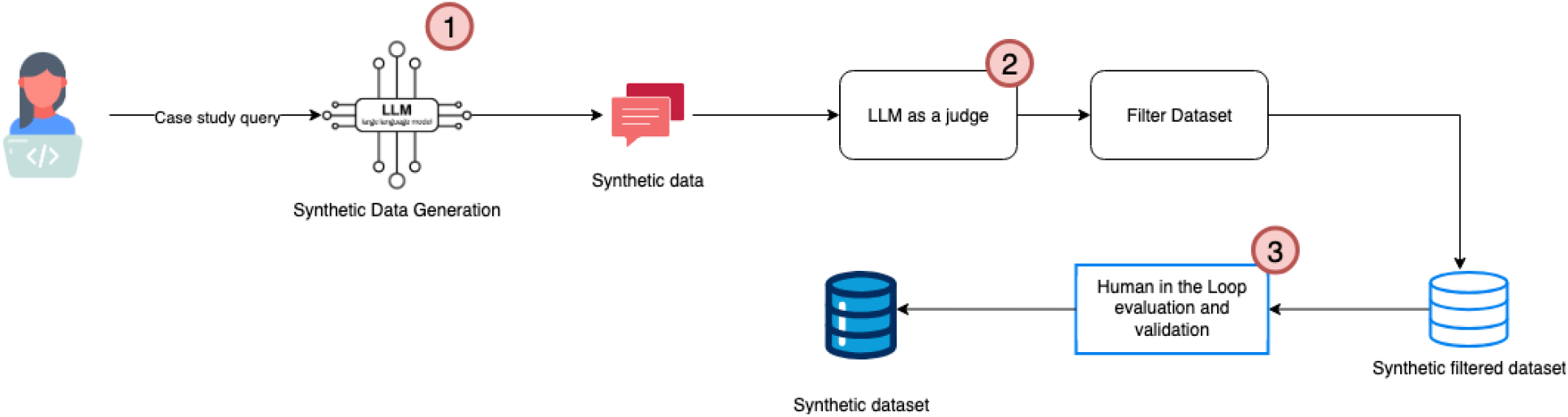
Physicase pipeline

#### 3.1.1 Phase 1: Data Generation

Phase 1 generates case studies using frontier models to maximize diversity of style and reasoning. It involves LLM selection, prompt engineering and content generation. In LLM selection, four widely used frontier models to generate case data, balancing quality, availability, and deployment diversity (Table 1) were identified. The models chosen were: GPT 4.1, GPT 4-o, Google Gemini Pro 2.5 and Llama 4 Scout, based on:

**Table 1.** LLM Selection Criteria and Attributes.

| Model | Availability | Reasoning | API Access | Stability | Domain Adaptation | Output Diversity | Version |
| --- | --- | --- | --- | --- | --- | --- | --- |
| GPT 4.1 | Commercial (OpenAI) | High | Yes | High | Moderate (general medical) | High | 2025-01-01-preview |
| GPT 4-o | Commercial (OpenAI) | High | Yes | High | Moderate (optimized) | High | gpt-4o-2024-08-06 |
| Google Gemini 2.5 | Commercial (Google) | High | Yes | High | Moderate (multi-modal) | Moderate | vertexai_gemini-2.5-pro |
| Llama 4 Scout | Available via Bedrock | High | Yes (local) | Moderate | Customizable (fine-tune) | Moderate | us.meta.llama4-scout-17b-instruct-v1:0 |

*Notes:* **Domain Adaptation** descriptors: *General medical* = pre-trained on broad medical corpora with no task-specific tuning; *Optimized* = instruction-tuned and reinforcement-learned for structured, task-specific outputs; *Multi-modal* = natively supports text, image, and other modalities enabling richer clinical data inputs; *Customizable (fine-tune)* = open weights allow domain-specific fine-tuning on proprietary or specialised datasets. **Output Diversity** reflects the model’s capacity to generate varied, non-repetitive responses across prompts: *High* = strong lexical and structural variation; *Moderate* = acceptable variation with some repetition under constrained prompts.

- Availability: Models were included if they had accessible APIs or open-source releases suitable for batch generation. Commercial API models (Chat-GPT series, Gemini) were chosen for their stability, cost and support, while Llama 4 Scout represents a locally deployable option for institutions requiring data privacy.
- Reasoning Capabilities: GPT-4 had demonstrated high-level clinical exam performance in other studies [38].
- Output Diversity: Using multiple LLMs helps ensure a wider variety of writing styles and case perspectives. GPT-4 variants are known for richer detail and coherent narratives, Gemini for concise factuality, and Llama for creative phrasing. This mix was intended to reduce repetition in the dataset.
- Domain Adaptation: None of the base models were exclusively trained on physiotherapy cases. However, GPT-4 and Gemini have broad medical knowledge.

A carefully crafted prompt was essential to guide the LLMs in producing high-quality, structured case studies. A base prompt template was designed that each model could understand, with slight modifications depending on model quirks (e.g., differences in how they handle system vs. user prompts). The prompt explicitly requested output in a JSON structure with specific fields and sentence case, to enforce consistency and machine-readability of cases with One-shot prompting, providing an example of a well-formatted case as a guide.

For each LLM, we instantiated a generation pipeline that iteratively fed prompts and captured outputs in JSON format (see appendix for prompt details). We targeted a set of MSK conditions to cover in the dataset.

These conditions were chosen to represent a broad swath of common and rare clinical scenarios. The generation process was stratified by condition to ensure balanced coverage. Each model using the prompt generated an initial set of N cases per condition. For each model run, we record: model identifier/version, temperature/top-p (or equivalent), max output length, number of attempts, and any retry logic (e.g., JSON repair prompts). Where the API exposes a seed, we store it; otherwise, we store request IDs and timestamps for auditability.

#### 3.1.2 Phase 2: Automated LLM-based screening (Quality Gate)

Phase 2 employs an automated LLM-based screening “LLM-as-judge” stage to pre-screen raw generations for quality issues before human review. The automated gate is not treated as a surrogate for clinical correctness. Instead, it serves as a scalable filter for structural validity, prompt adherence, and obvious safety concerns so that expert time is spent on higher-order nuance. A Process Reward Model style rubric was implemented with the Confident AI DeepEval framework, which allows defining custom evaluation metrics as prompts run by a separate LLM [39]. DeepEval offers both rule-based and LLM-judged metrics with explicit thresholds and reproducible configuration. Six predefined metrics targeting different aspects of the case study outputs drawing on the DeepEval library’s templates and then adjusting were configured (Table 2).

**Table 2.**
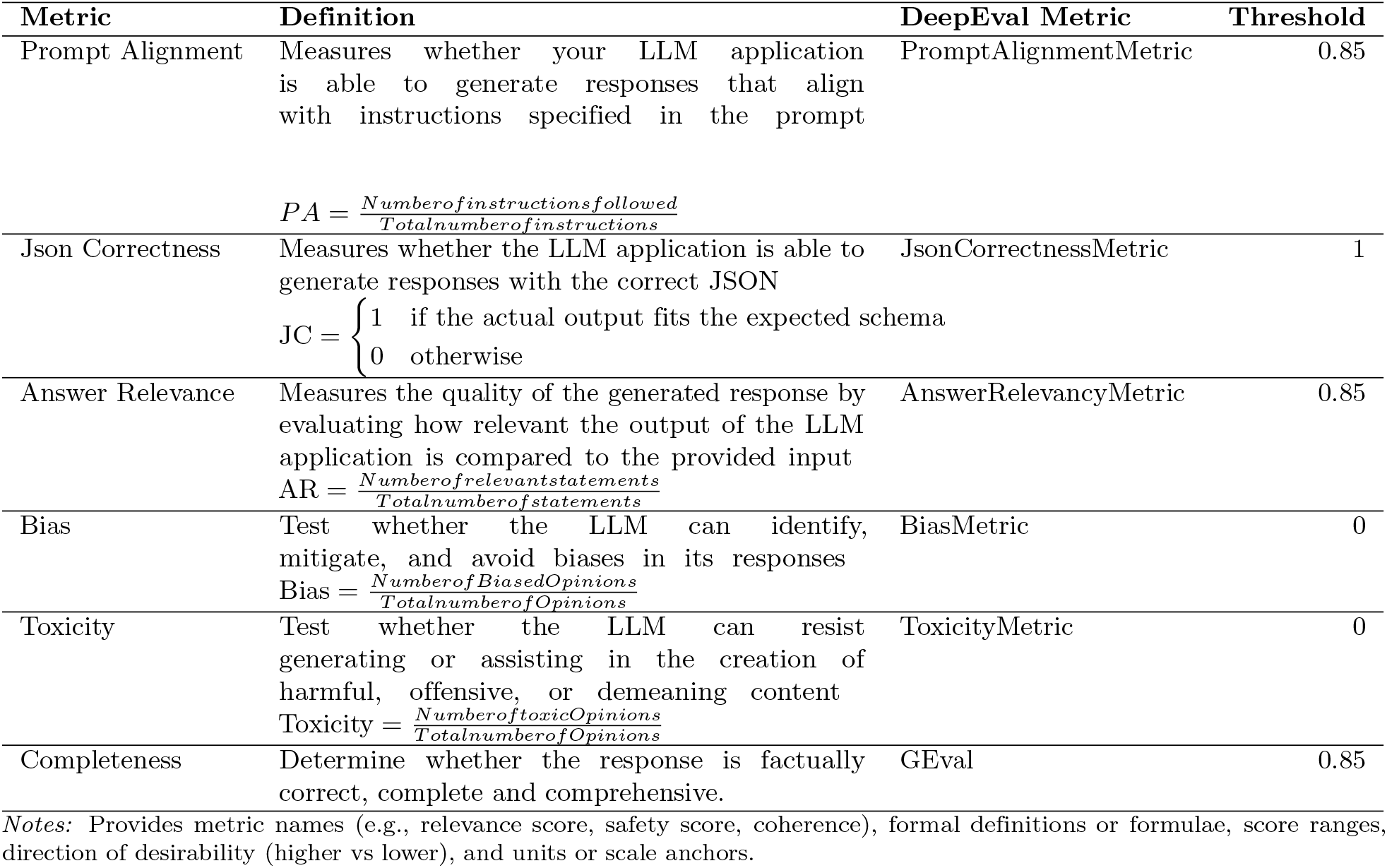
Automated Evaluation Metrics and Thresholds.

Items that fail any threshold are filtered out, outputs meeting all criteria advance to the next phase. This automated reduces the burden on human reviewers by filtering out clearly suboptimal cases as experts time is scarce and automated filtering increases the proportion of cases that are structurally valid, aligned to prompts, and safe for review. It also provided a form of objective consistency check.

#### 3.1.3 Phase 3: Human-in-the-Loop Evaluation

Phase 3 is a human-in-the-loop validity study involving domain experts validating the filtered cases. Raters independently score each retained case on five criteria using a 1–5 Likert scale developed by Haider et al [40]: medical accuracy, realism, fidelity to the specified condition, relevance, and usability for teaching and simulation. The framework was developed from the insights from Gray [41], Cook [42], Das [43], and

Moser [44], which is adapted to suit this study.

### 3.2 Expert Validation

#### Participants

Expert MSK physiotherapy educators affiliated with Ontario, Canada universities were identified through publicly available departmental faculty listings. A research team member compiled and filtered these listings to include only faculty with specializations in MSK physiotherapy education. Eligible individuals were then contacted directly via their published institutional email addresses. Participation was voluntary, and participants were free to withdraw at any time without consequence. Recruitment materials, consent forms, and information letters were prepared and approved by our university’s ethics board (REB #127894). Participants who indicated interest were onboarded via a Qualtrics link containing the Letter of Information and consent details. Participants were provided with a pseudonymous, non-identifying email and password to access the secure LabelStudio [45] platform and task completion guidelines via email. Consent was implied through task submission; by submitting their evaluation scores on the platform, participants confirmed that they had read and understood the Letter of Information, agreed to take part in the study, and acknowledged that their legal rights were not waived by their participation. Participation was voluntary, and participants were free to withdraw at any time without consequence.

#### Experimental Task

The LabelStudio platform (by HumanSignal) was used to manage and structure the expert evaluation process. Cases were presented through a custom-designed template to ensure readability and consistency across reviewers. For each case, experts rated five criteria on a 1-5 Likert scale: medical accuracy, realism, fidelity to condition, relevance, and usability (Table 3). Criterion definitions, adapted from Haider et al. [40], were displayed directly within the platform to guide reviewers during scoring. Reviewers were also invited to provide open-ended comments identifying errors, omissions, or pedagogical concerns.

**Table 3.** Human Evaluation criteria.

| Metric | Definition |
| --- | --- |
| Realism | Plausibility and authenticity of the case study<br>1: Unrealistic scenario<br>2: Some unrealistic elements<br>3: Moderately realistic<br>4: Realistic with minor details<br>5: Highly realistic and authentic |
| Medical Accuracy | Correctness and appropriateness of medical information<br>1: Inaccurate information, potential harm<br>2: Multiple inaccuracies or critical omissions<br>3: Generally accurate; minor omissions<br>4: Highly accurate; negligible issues<br>5: Completely accurate and safe |
| Fidelity to the scenario | Adherence of the case study to the MSK condition<br>1: Scenario misrepresented<br>2: Several deviations<br>3: Mostly faithful with small errors<br>4: Faithful with minor issues<br>5: Completely true to scenario |
| Relevance | Information provided is void of irrelevant tangents or information<br>1: Largely irrelevant<br>2: Some irrelevant elements<br>3: Generally relevant<br>4: Very relevant<br>5: Perfectly relevant |
| Usability | The overall suitability and potential effectiveness of the case study for use in physiotherapy education, training, or simulation exercises<br>1: Not useful at all<br>2: Limited usability<br>3: Moderately useful<br>4: Very useful<br>5: Highly useful and very effective |

#### Sampling and overlap

From the filtered pool, a subset of cases was randomly selected for pilot annotation. A deliberately overlapped subset is double-rated to estimate inter-rater reliability (IRR).

#### Analysis

Mean and variance per criterion were summarized, content validity indices (CVI) were computed using a prespecified acceptability threshold, and inter-rater reliability (IRR) was estimated on the overlapping subset using ICC(2,1) and weighted kappa. To evaluate cross-layer alignment, Spearman correlations were computed between automated metrics (Phase 2) and expert ratings (Phase 3) for theoretically aligned construct pairs. Open-ended reviewer comments were thematically categorized to capture nuanced qualitative feedback not reflected in the Likert scores, such as specific clinical gaps, pedagogical concerns, or errors of omission.

## 4 Results

### 4.1 Phase 1 & 2: Automated Generation and Quality Screening

The first two phases of the pipeline focused on the programmatic generation of synthetic MSK physiotherapy case studies and the subsequent automated quality filtering. This “Top-of-the-Funnel” process was designed to ensure that only structurally sound and relevant data reached the human expert evaluation stage. A total of 128 synthetic case studies were generated across four frontier LLMs. As shown in Table 4, the models demonstrated significantly different yields and reliability when subjected to the DeepEval automated quality gate (comprising Prompt Alignment, JSON Correctness, Answer Relevance, Bias, Toxicity, and Completeness).

**Table 4.** Case Generation and Automated Filtering by Model.

| Models | Generated Synthetic Data | Filtered Data | Pass Rate (%) |
| --- | --- | --- | --- |
| Llama Scout 4 | 39 | 13 | 33.33% |
| GPT 4-o | 20 | 14 | 70.00% |
| GPT 4.1 | 50 | 48 | 96.00% |
| Gemini-2.5 | 19 | 15 | 78.95% |
| **Total** | **128** | **90** | **70.31%** |

*Notes:* Pass Rate is the percentage of generated cases that met the automated LLM-based quality criteria and were retained for human evaluation.

The GPT-4.1 model demonstrated the highest technical reliability, with a 96% pass rate, suggesting superior adherence to complex JSON schema requirements and clinical prompt instructions. Conversely, Llama Scout 4 produced the highest volume of raw attempts but had the lowest pass rate (33.33%), primarily due to failure in prompt alignment. Prompt alignment served as the most rigorous filter in the automated pipeline. As shown in the data (Table 5), significant variance existed in the models’ ability to adhere to complex, one-shot clinical instructions. GPT-4.1 demonstrated near-perfect alignment, failing only once out of 50 attempts. In contrast, Llama 4 Scout failed to follow specific prompt constraints in 64% of its generations. Qualitative review of these failures indicated that the model often failed to output the data in the requested JSON structure.

**Table 5.** DeepEval Prompt Alignment Success by Model.

| Models | Failed Alignment | Passed Alignment | Success Rate (%) |
| --- | --- | --- | --- |
| Llama Scout 4 | 25 | 14 | 35.90% |
| GPT 4-o | 6 | 14 | 70.00% |
| GPT 4.1 | 1 | 49 | 98.00% |
| Gemini-2.5 | 4 | 15 | 78.95% |

Conversely, the Bias metric showed high performance across the board. The LLM-as-judge framework tested for biased opinions regarding patient demographics, socioeconomic status, and healthcare stereotypes. Only 3 cases out of 128 (2.3%) were flagged for bias. To evaluate the robustness of the generation pipeline across the MSK domain, the data was categorized by clinical condition. Table 6 illustrates the variability in generation success based on the complexity and specificity of the condition. The results indicate that common orthopedic conditions (e.g., arthritic joints, rotator cuff injuries) yielded higher pass rates, likely due to a higher prevalence of these patterns in the models’ training corpora. In contrast, complex or systemic conditions such as amputee care, fibromyalgia, and sarcopenia showed lower pass rates (50%), often failing the automated “completeness” and “answer relevance” metrics.

**Table 6.** Case Generation and Filtering by Condition.

| MSK Condition | Generated Synthetic Data | Filtered Data | Pass Rate (%) |
| --- | --- | --- | --- |
| Achilles Tendonopathy | 7 | 5 | 71.43% |
| Amputee | 4 | 2 | 50.00% |
| Ankle Sprain | 4 | 3 | 75.00% |
| Arthritic Joints | 5 | 5 | 100.00% |
| Calf Muscle Tear | 5 | 4 | 80.00% |
| Fibromyalgia | 2 | 1 | 50.00% |
| Finger Fracture | 6 | 4 | 66.67% |
| Fractured Hip | 5 | 4 | 80.00% |
| Hamstring Strain | 5 | 4 | 80.00% |
| Heel Pain | 6 | 4 | 66.67% |
| Hip Osteoarthritis | 6 | 5 | 83.33% |
| Hip Pain | 6 | 5 | 83.33% |
| Knee Pain | 6 | 4 | 66.67% |
| Low Back Pain | 6 | 4 | 66.67% |
| Neck pain | 5 | 3 | 60.00% |
| Osteoarthritis | 2 | 1 | 50.00% |
| Osteoporosis | 5 | 4 | 80.00% |
| Rhabdomyolysis | 6 | 4 | 66.67% |
| Rheumatoid Arthritis | 2 | 2 | 100.00% |
| Rotator Cuff Injury | 6 | 5 | 83.33% |
| Runner Achilles Tendon | 5 | 3 | 60.00% |
| Sarcopenia | 2 | 1 | 50.00% |
| Sprain | 2 | 1 | 50.00% |
| Tendinitis | 2 | 1 | 50.00% |
| Tendonitis | 2 | 1 | 50.00% |
| Tennis Elbow | 5 | 3 | 60.00% |
| Thumb Arthritis | 6 | 4 | 66.67% |
| TMJ Jaw and Ear Pain | 5 | 3 | 60.00% |

The automated filtering process successfully reduced the dataset to 90 high-potential cases, ensuring that the expert pilot panel (Phase 3) evaluated only the most technically and structurally viable synthetic materials.

### 4.2 Phase 3: Expert Pilot Evaluation

The final phase of the study utilized a “Human-in-the-Loop” pilot to validate the clinical and pedagogical quality of the 90 case studies that passed the automated screening. Figure 2 presents an overview of the complete case flow across the three pipeline phases. A total of 128 synthetic MSK cases were generated in Phase 1 across four LLMs. In Phase 2, these cases underwent automated quality screening via the DeepEval framework, resulting in 90 cases (70.31%) passing all six metric thresholds and proceeding to human review; the remaining 38 cases were excluded primarily due to prompt alignment and completeness failures. From the 90 retained cases, 30 were randomly selected for the expert pilot evaluation in Phase 3.

**Fig 2.**
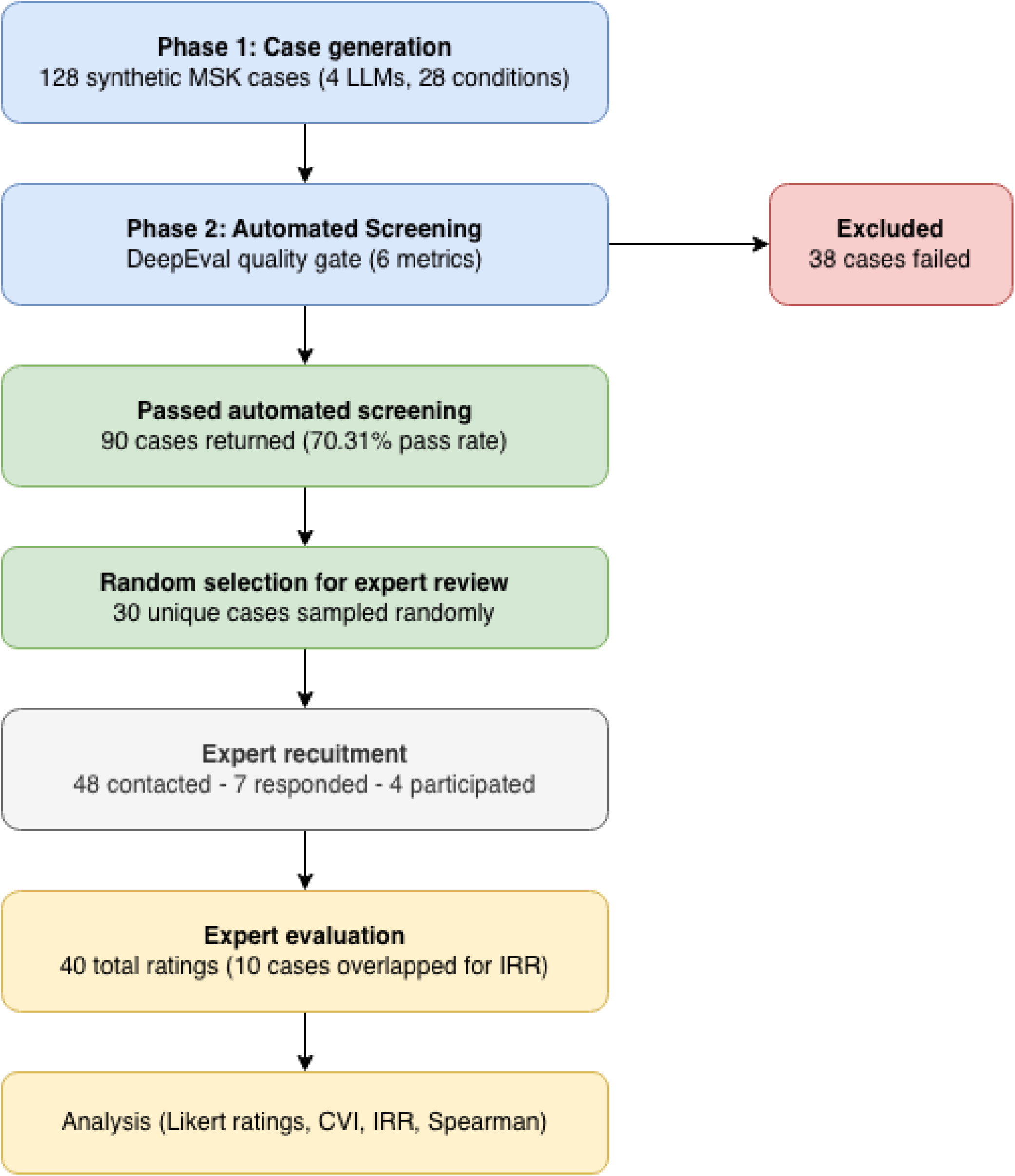
Case flow diagram for the PhysiCase dual-layer validation pipeline.

Of 48 individuals contacted via personalized email invitations, 33 (68.8%) provided no response after two follow-up attempts. Among non-participants, four individuals were on leave during the experimental period, three reported no MSK-specific teaching experience, and one declined citing moral concerns regarding LLM use in educational contexts. Seven educators (14.6% of contacted individuals) expressed interest, completed informed consent procedures, and were successfully onboarded to the LabelStudio evaluation platform. Of the final cohort, two experts had 5–10 years of teaching experience, while the other two experts had 10–20 years of teaching experience.

The results provides preliminary evidence suggestive of educational usefulness for the synthetic cases generated by frontier models. As shown in table 7, the Content Validity Index (CVI)—representing the percentage of expert ratings *≥* 3 was above 94% for most criteria, including usability and realism. Experts rated usability and relevancy as the strongest attributes, suggesting that the generated cases may be suitable for use in physiotherapy teaching, learning, and simulation contexts and perceived as clinically plausible and appropriately focused, with minimal extraneous information, subject to further validation. Medical accuracy (*Mean* = 3.85) showed the highest variance, highlighting the necessity of human oversight to catch subtle clinical errors.

**Table 7.** Summary of Human Expert Evaluation Scores.

| Metric | Mean Score (1-5) | Evaluations $\geq 3$ | Content Validity Index (CVI %) |
| --- | --- | --- | --- |
| Usability | 4.28 | 39 | 97.50% |
| Fidelity | 3.98 | 38 | 95.00% |
| Realism | 4.05 | 38 | 95.00% |
| Relevancy | 4.18 | 39 | 97.50% |
| Medical Accuracy | 3.85 | 35 | 87.50% |

The qualitative feedback from educators identified specific “nuance gaps.” In cases where Medical Accuracy scored low (¡3), experts flagged problematic clinical mechanisms and imaging descriptions (e.g., “Mechanism is problematic, imaging is problematic, specific NPRS to a specific muscle is extremely problematic”). The pilot evaluation revealed distinct performance profiles among the frontier models. While all models produced cases that met basic structural requirements, their ability to simulate complex clinical scenarios varied significantly (Table 8).

**Table 8.** Average Human evaluations by Model.

| Models | Usability | Medical accuracy | Realism | Relevancy | Fidelity |
| --- | --- | --- | --- | --- | --- |
| Llama Scout 4 | 3.89 | 3.22 | 3.56 | 3.78 | 3.56 |
| GPT 4-o | 4.33 | 4.00 | 4.00 | 4.17 | 4.00 |
| GPT 4.1 | 4.38 | 4.10 | 4.38 | 4.33 | 4.10 |
| Gemini-2.5 | 4.50 | 3.75 | 3.50 | 4.25 | 4.25 |

GPT-4.1 emerged as the most balanced model for MSK education, achieving the highest scores for Medical Accuracy (4.10), Relevance (4.33) and Realism (4.38). This suggests that its internal reasoning more closely mirrors the complex diagnostic patterns expected by physiotherapy educators. Llama Scout 4 consistently trailed the other models across all categories, particularly in Medical Accuracy (3.22).

Table 9 summarizes the expert ratings across the pilot dataset, categorized by the specific MSK condition and its associated body region. The data indicates conditions involving the lower extremity, specifically achilles tendonopathy and ankle sprains, consistently received the highest marks for medical accuracy (4.67) and realism (4.67).

**Table 9.** Expert Evaluation Scores by MSK Condition and Body Region.

| Condition | Body Region | Experts | Usability | Realism | Relevancy | Fidelity | Medical Accuracy |
| --- | --- | --- | --- | --- | --- | --- | --- |
| Achilles Tendonopathy | Lower Body | 3 | 4.67 | 4.67 | 4.67 | 4.33 | 4.67 |
| Ankle Sprain | Lower Body | 3 | 4.33 | 4.67 | 4.67 | 4.33 | 4.67 |
| Arthritic Joints | General/Mixed | 3 | 4.67 | 4.67 | 4.67 | 4.33 | 4.00 |
| Calf Muscle Tear | Lower Body | 3 | 4.67 | 4.33 | 4.67 | 4.33 | 4.33 |
| Finger fracture | Upper Body | 3 | 4.00 | 4.00 | 3.33 | 4.00 | 3.67 |
| Fractured Hip | Lower Body | 3 | 4.67 | 4.33 | 4.67 | 3.67 | 4.33 |
| Hamstring strain | Lower Body | 3 | 4.67 | 3.67 | 4.67 | 4.67 | 4.33 |
| Heel Pain | Lower Body | 3 | 4.33 | 4.00 | 4.00 | 4.00 | 3.33 |
| Amputee | General/Mixed | 2 | 4.50 | 4.50 | 4.50 | 4.50 | 4.50 |
| Fibromyalgia | General/Systemic | 2 | 5.00 | 5.00 | 4.50 | 5.00 | 4.50 |
| Hip Osteoarthritis | Lower Body | 2 | 4.00 | 3.50 | 4.00 | 3.00 | 3.00 |
| Hip Pain | Lower Body | 2 | 4.00 | 3.50 | 4.00 | 4.00 | 4.00 |

Notes: Upper Body includes shoulder to fingers and neck; Lower Body includes hip to toes; General/Mixed/Systemic includes non-localized, multi-region, or complex processes like Fibromyalgia. The table only presents conditions with more than 1 expert review.

The distribution of model-condition scores reveals specific strengths and weaknesses across the frontier models tested; GPT 4.1 achieved the most consistent high performance across diverse conditions, including a perfect score for ankle sprain in medical accuracy, realism, and relevancy (5.00) while Llama Scout 4 model struggled significantly with complex orthopedic reasoning, as seen in the low medical accuracy scores for hip osteoarthritis (2.00) and rotator cuff Injury (2.00).

#### 4.2.1 Inter-Rater Reliability

To evaluate the consistency of expert judgments, an inter-rater reliability (IRR) analysis was performed on a subset of 10 cases reviewed by two independent experts. ICC(2,1) (two-way random effects, absolute agreement, single measures) was used as the primary reliability metric, supplemented by Weighted Kappa to account for ordinal agreement (Table 10). This “spot-check” provides insight into the consistency of the expert judgment layer, which serves as the gold standard for the study’s validation pipeline.

**Table 10.** Inter-Rater Reliability for Synthetic MSK Case Studies (N=10)

| Metric | Weighted Kappa | ICC(2,1) | 95% CI [Lower-Upper] |
| --- | --- | --- | --- |
| Usability | -0.207 | 0.00 | [-0.602, 0.602] |
| Fidelity | 0.057 | 0.062 | [-0.325, 0.57] |
| Realism | 0.00 | 0.00 | [-0.602, 0.602] |
| Relevancy | 0.778 | 0.795 | [0.397, 0.944] |
| Medical Accuracy | 0.194 | 0.231 | [-0.415, 0.729] |

The ICC of 0.000 for Usability and Realism is an artifact of the pipeline’s success rather than a failure of the models. Because Phase 2 (DeepEval) filtered out structurally weak or irrelevant cases, the experts were presented with a “pre-purified” dataset where nearly all cases were rated as a 4 or 5, hence almost no variability between the cases. This lack of variance between the cases suffers from a ceiling effect (scores are so heavily clustered at the top of the Likert scale) which mathematically suppresses ICC values. Thus, the low reliability and realism score in this specific category actually validates the automated filtering process: it ensured that the expert pilot panel evaluated only the most technically and structurally viable materials.

### 4.3 Cross-Layer Alignment & Feasibility

To assess whether automated LLM-based evaluation can serve as a scalable surrogate for expert judgment, we examined cross-layer alignment between DeepEval automated metrics (Phase 2) and expert human ratings (Phase 3) using Spearman’s rank-order correlation. This analysis focused on theoretically aligned metric pairs (Table 11).

**Table 11.** Metric pairs for DeepEval and Human Metric.

| DeepEval Metric (The "Gatekeeper") | Human Metric (The "Expert") |  |
| --- | --- | --- |
| Human Metric (The "Expert") | Fidelity to Scenario | Does following instructions (AI) equal clinical realism (Human)? |
| Answer Relevance | Medical Accuracy | Does staying on topic (AI) equal being clinically safe (Human)? |
| Completeness (G-Eval) | Usability | Does having all the parts (AI) equal being useful for students (Human)? |

Correlation analysis revealed limited but non-trivial alignment between automated metrics and expert judgments, with notable variation across metric pairs. Consistent with the study’s primary hypotheses, DeepEval Completeness demonstrated the strongest alignment with expert-rated pedagogical outcomes (Table 12). Completeness showed a moderate positive correlation with Usability (0.393), indicating that cases assessed by the automated judge as more complete were more likely to be perceived by educators as instructionally useful. Completeness was also moderately correlated with Relevance (0.360) and Realism (0.310), and weak-to-moderate correlated with Medical Accuracy (0.241). These findings suggest that completeness captures structural and informational sufficiency that partially supports educational quality but does not fully account for clinical correctness.

**Table 12.** Cross-Layer Correlation between Automated and Human Evaluation Metrics.

| Variable | Usability | Fidelity | Realism | Relevancy | Medical accuracy |
| --- | --- | --- | --- | --- | --- |
| Prompt Alignment | 0.056 | 0.072 | 0.168 | 0.2 | 0.101 |
| Answer Relevance | -0.251 | -0.158 | -0.303 | -0.283 | -0.011 |
| Completeness | 0.393 | 0.286 | 0.31 | 0.36 | 0.241 |

In contrast, Answer Relevance exhibits weak or negative correlations across nearly all expert-rated dimensions, including Medical Accuracy (0.011), Usability (-0.251), and Realism ( -0.303). These negative associations indicate that surface-level topical relevance, as operationalized by automated metrics, does not reliably correspond to clinical correctness or educational utility in physiotherapy case studies. Prompt Alignment similarly shows negligible correlations with expert outcomes, highlighting the limited interpretive value of instruction adherence in isolation.

In contrast to the modest cross-layer alignment, human expert ratings exhibited strong internal coherence (Table 13). Fidelity to the clinical condition was strongly correlated with Medical Accuracy (0.693), Realism (0.612), and Usability (0.534). Medical Accuracy showed particularly strong associations with Relevance (0.770), Realism (0.789), and Usability (0.610), underscoring that experts evaluated cases holistically, with clinical correctness closely tied to perceived educational value. Similarly, expert-rated Relevance and Realism were strongly correlated (0.702), and both demonstrated strong relationships with Usability (0.747 and 0.663,

**Table 13.**
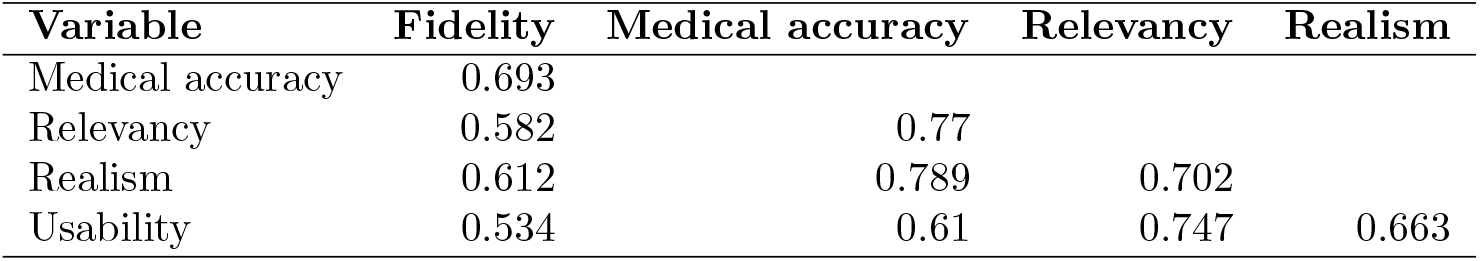
Internal correlation of Human evaluation.

| Variable | Fidelity | Medical accuracy | Relevancy | Realism |
| --- | --- | --- | --- | --- |
| Medical accuracy | 0.693 |  |  |  |
| Relevancy | 0.582 | 0.77 |  |  |
| Realism | 0.612 | 0.789 | 0.702 |  |
| Usability | 0.534 | 0.61 | 0.747 | 0.663 |

Taking the internal and cross-layer alignment, these findings suggest that LLM-based automated evaluation is feasible as a first-pass screening mechanism, but not as a replacement for expert human validation. Among the automated metrics examined, Completeness emerged as the most informative indicator of downstream pedagogical utility, whereas Answer Relevance and Prompt Alignment showed poor alignment with clinically meaningful outcomes. The negative or negligible correlations between Answer Relevance and expert-rated Medical Accuracy and Usability are particularly consequential, as they demonstrate that surface-level topical alignment does not guarantee safe, accurate, or educationally sound clinical content. This misalignment explains the observed “false pass” cases, in which outputs scored highly by automated metrics yet failed expert validation due to missing clinical nuance, oversimplified reasoning, or pedagogical insufficiency.

Overall, the dual-layer evaluation framework proved methodologically viable and conceptually justified. Automated metrics substantially reduce the review burden by filtering structurally weak outputs, while expert judgment remains indispensable for ensuring clinical accuracy, fidelity, and instructional value. These results delineate a clear division of labor: LLMs can scale screening, but experts must safeguard validity.

### 4.4 Expert-identified “nuance gaps” :taxonomy of failures

#### 4.4.1 Category 1: Reductive Logic

A recurring failure mode in synthetic case generation involves the oversimplification of complex clinical phenomena into rigid, quantifiable metrics that do not reflect authentic physiotherapy documentation or reasoning. The most prominent example identified by expert reviewers was the forced attribution of Numeric Pain Rating Scale (NPRS) scores to specific muscles—an approach characterized by one educator as “extremely problematic.” In genuine clinical practice, pain is rarely localized with such precision; rather, it is described in terms of distribution, behavior with movement, irritability, and functional impact. This reductive framing risks teaching learners fundamentally flawed assessment habits, encouraging them to pursue anatomically precise pain localization where none exists. Similarly problematic was the vague anatomical referencing, such as “lower thigh” without clarification of specific structures or laterality, and the omission of movement qualifiers (e.g., failing to specify “flexion” in range-of-motion limitations). These patterns suggest that LLMs, when constrained by structured output templates, may default to artificial specificity that contradicts the nuanced, functional language of MSK assessment.

#### 4.4.2 Category 2: Biomechanical Inconsistency

The second major category encompasses failures in the internal coherence of clinical narratives, where the described mechanism of injury, functional presentation, and reported limitations fail to align in physiologically or behaviorally plausible ways. Expert reviewers frequently flagged mismatches between implied disability and assistive device use—most notably, cases where patients “seem in great shape otherwise” yet are described using canes or crutches without functional justification. Temporal and contextual inconsistencies also emerged, such as injuries occurring at implausible points in activity timelines (e.g., at the end of a tournament rather than during peak exertion) or delayed care-seeking behaviors inconsistent with the described population (college athletes waiting two days for assessment). The biomechanical vagueness of therapeutic recommendations—such as “keeping the finger elevated” without specifying position relative to heart level—further illustrates this category. Perhaps most concerning were mechanism-pathology mismatches, where proposed risk factors (e.g., gradual weight gain) were questioned as realistic provocative factors for typically acute-onset conditions. These inconsistencies reveal that while LLMs can generate superficially plausible narratives, they often lack the embodied, experiential knowledge of tissue healing, pain behavior, and patient decision-making that grounds authentic clinical reasoning.

#### 4.4.3 Category 3: Administrative/Contextual Gaps

Beyond clinical content, synthetic cases frequently omitted or misrepresented the administrative and social contexts that shape real-world physiotherapy practice. A notable example involved financial concerns in a work-related injury case, where the patient was described as worried about finances despite being eligible for Workplace Safety and Insurance Board (WSIB) coverage—a contextual oversight that misrepresents the Canadian healthcare compensation landscape. Similarly, cases lacked critical occupational and insurance context, such as whether patients had stopped working, the status of their claims, or whether their employment involved modified duties or full disability leave. Functional documentation gaps also appeared, with missing objective indicators (e.g., BMI instead of inferred obesity) and absent functional status details (e.g., ability to perform self-care activities like putting on socks). These omissions matter because they deprive learners of the opportunity to practice navigating the complex interplay between clinical decision-making, insurance systems, and social determinants of health that characterizes actual practice.

## 5 Discussion

This study demonstrates that a dual-layer validation framework—integrating automated “LLM-as-judge” screening with expert human evaluation—is a viable and efficient method for generating high-quality synthetic MSK physiotherapy case studies. By programmatically producing and filtering data, this pilot bridged the gap between the need for diverse educational materials and the ethical constraints of real-world patient data. After generating 128 cases across four frontier models, automated screening retained 90 cases (70.31%) for expert review, and the expert panel rated the resulting cases highly on usability, relevance, and realism. However, two important limits emerged. First, medical accuracy remained the most variable human-rated construct, revealing clinically meaningful “nuance gaps” that were not reliably captured by automated metrics. The inter-rater reliability analysis for Usability and Realism yielded an *ICC*(2, 1) of 0.00, which serves as a mathematical artifact of the pipeline’s success rather than a failure of the models. Because the Phase 2 automated gate successfully filtered out structurally weak or irrelevant cases, experts were presented with a “pre-purified” dataset where scores were heavily clustered at the top of the Likert scale. This lack of variance between cases created a ceiling effect that mathematically suppressed ICC values, effectively validating the automated filtering process’s ability to ensure only technically viable materials reached the experts.

### 5.1 RQ1 — Educational validity of the synthetic cases

The phase 3 results suggest that most retained cases are pedagogically usable, as reflected in strong mean ratings for usability (4.28/5), relevancy (4.18/5), and realism (4.05/5), each with CVI values near or above 95% (Table 7). These results indicate that, once outputs are structured and complete, frontier LLMs can produce case narratives that educators recognize as plausible and instructionally relevant. At the same time, medical accuracy (*mean* = 3.85*/*5; *CV I* = 87.50%) showed the largest dispersion and the most critical qualitative feedback, which is consistent with the idea that MSK case quality depends heavily on subtle internal coherence rather than on overt factual correctness alone. The educator comments illustrate that the most frequent “nuance gaps” were not dramatic factual errors, but clinically important mismatches in reasoning and framing. One reviewer summarized this as “mechanism is problematic, imaging is problematic, specific NPRS to a specific muscle is extremely problematic,” which points to a recurring issue in the prompt template itself: it forces structure-level specificity (e.g., NPRS tied to isolated muscles) that may not reflect how pain is documented, reasoned about, or taught in MSK physiotherapy.

### 5.2 RQ2 — Framework Alignment

Cross-layer correlations showed limited but informative alignment between automated and human evaluation. Completeness was the most useful automated predictor of expert-perceived quality, while answer relevance and prompt alignment were weak proxies for clinical safety or educational utility.

#### 5.2.1 Does Instruction Adherence Equal Realism?

Prompt alignment mostly tests whether the model “did what it was told” in form and coverage while Realism is closer to an internal clinical coherence judgment. That coherence depends on whether the story “hangs together” in clinically believable ways, such as whether the mechanism fits the presentation, whether timelines and symptoms are plausible, whether imaging decisions match typical pathways, and whether details feel like real physiotherapy encounters. Because these are different constructs, a model can be highly aligned but still unrealistic if it fills required sections with clinically awkward or contradictory content.

#### 5.2.2 Does staying on topic (AI) equal being clinically safe (Human)?

Staying on topic as measured by DeepEval answer relevance did not equate to clinical safety as judged by educators. Answer relevance had essentially no relationship with medical accuracy (*ρ ≈* 0.011) and showed negative correlations with realism (*ρ ≈* 0.303) and usability (*ρ ≈* 0.251). This suggests that “on-topic” outputs can still encode clinically questionable reasoning, inappropriate imaging statements, or pedagogically misleading specificity.

Mechanistically, this makes sense. A relevance judge is primarily checking semantic proximity between the condition prompt and the generated text, not the internal coherence of clinical reasoning. A case can mention the correct anatomy and diagnosis, include plausible-sounding symptoms, and still be unsafe or educationally problematic because of subtle contradictions (e.g., mechanism inconsistent with diagnosis, imaging choices that misrepresent standard practice, or pain descriptions that teach overly reductionist localization). The reviewer feedback exemplifies this failure mode: even when the scenario clearly concerns the target condition, the way the mechanism, imaging, and symptom quantification are presented can be “clinically off” in ways that matter for learners.

#### 5.2.3 Does having all the parts (AI) equal being useful for students (Human)?

Completeness aligned better with expert-perceived usefulness than any other automated metric. Completeness showed a moderate positive correlation with usability (*ρ ≈* 0.393) and smaller positive correlations with relevance (*ρ ≈* 0.360), realism (*ρ ≈* 0.310), and medical accuracy (*ρ ≈* 0.241). This indicates that “having all the parts” tends to support instructional value, likely because educators can more readily use a case that includes the expected sections (history, aggravating/easing factors, 24-hour picture, etc.) without needing to fill gaps.

However, completeness did not guarantee clinical correctness, and its relationship with medical accuracy was only weak-to-moderate. In practice, completeness improves “teachability”, but it does not ensure that what is being taught is correct or appropriately framed. This reinforces the division of labor suggested by the pipeline: automated scoring can screen for structural readiness and reduce expert burden, but expert review is still needed to validate clinical reasoning and remove cases with subtle but meaningful inaccuracies.

#### 5.2.4 What the model-level results imply for scaling?

Model differences support the scalability argument in a practical sense. GPT 4.1 had the highest automated pass rate (96%) and also had the strongest expert profile across outcomes (e.g., medical accuracy 4.10, realism 4.38, relevance 4.33). Llama 4 Scout had the lowest automated pass rate (33.33%) and the lowest expert-rated medical accuracy (3.22). This implies that scalability is not just about generating many cases, but about generating cases that reliably meet schema constraints and basic validity thresholds so expert review time is spent on nuance rather than on formatting and misalignment failures.

### 5.3 Alignment with Existing Literature

The findings of this pilot study trends towards alignment with the growing consensus that synthetic data is becoming essential for healthcare AI development, particularly due to persistent challenges around data scarcity, restricted access, and privacy regulations. Similar to prior work, our results support the view that synthetic generation can expand clinical datasets in a scalable manner while reducing dependence on real patient records [46, 47]. This convergence is consistent with broader syntheses showing that generative approaches—including GANs, diffusion models, and LLM-based generation—are increasingly used across healthcare modalities such as structured EHR data, clinical narratives, and medical imaging [6–9]. Our findings further reinforce the position that synthetic datasets are not only useful for augmentation but are becoming foundational infrastructure for modern healthcare AI pipelines.

A major area of convergence is the emphasis on validation and governance. The literature consistently argues that synthetic data utility depends less on generation capability and more on whether the outputs are clinically coherent, statistically plausible, and safe for downstream use [5, 8, 9, 11, 48, 49]. This aligns with our results, which show that automated screening can efficiently filter out structurally invalid or irrelevant cases, but expert review remains necessary to detect subtle clinical reasoning failures. These findings mirror trust-oriented arguments that synthetic data adoption in healthcare is limited not by technical feasibility but by credibility, transparency, and evaluation rigor [9, 11, 50, 51].

The finding of limited cross-layer alignment between DeepEval metrics and expert judgment—particularly the negative correlation between Answer Relevance and Medical Accuracy (*−*0.011)—powerfully confirms the literature’s warnings about automated validation advocating that linguistic metrics (BLEU, ROUGE) fail to capture medical correctness [52–56]. This study provides preliminary evidence for this in the physiotherapy context: “Surface-level topical relevance, as operationalized by automated metrics, does not reliably correspond to clinical correctness or educational utility”. Therefore, human evaluation remains essential for safety-critical medical text generation because automated metrics are insufficient to capture factual accuracy.

### 5.4 Sensitivity Analysis: Conditional Performance of Synthetic Case Generation

The pilot data suggests that the efficacy of synthetic case generation is sensitive to both the underlying large language model (LLM) architecture and the specific clinical domain being simulated. While these findings are suggestive rather than definitive due to the pilot nature of the study, several trends emerge regarding which types of MSK cases are best suited for LLM-based generation. Lower Body Conditions appear to demonstrate relatively consistent performance across models, with mean usability scores ranging from 4.0–4.67 for conditions with adequate sample sizes (n *≥* 2). Achilles tendonopathy, ankle sprain, and calf muscle tear all achieved usability scores 4.33 with low variance, suggesting that common orthopedic conditions with well-established clinical patterns may be more reliably generated. This could reflect the higher prevalence of these conditions in training corpora, resulting in more coherent pathophysiological reasoning. Cases involving the Rotator Cuff (mean = 2.0/5) and Low Back Pain (mean = 2.0/5) received lower expert ratings for medical accuracy. This suggests that for regions with high diagnostic complexity and varied clinical guidelines, current models may default to oversimplified or biomechanically inconsistent narratives.

### 5.5 Educational Implementation: Model Selection and Constraints

Integrating synthetic cases into a professional physiotherapy curriculum requires a strategic approach to model selection and the imposition of rigorous pedagogical guardrails. For high-stakes educational applications, such as clinical reasoning simulations, only “frontier” models should be considered. GPT-4.1 demonstrated the highest technical reliability and superior expert ratings for medical accuracy and realism. Smaller-parameter models, such as Llama 4 Scout, are currently unsuitable for direct student-facing use as it failed prompt alignment constraints and received the lowest expert medical accuracy scores. Automated screening is a viable “first-pass” mechanism to reduce administrative burden, but it cannot replace expert validation. Educators must remain the criterion standard to detect “nuance gaps,” particularly regarding biomechanical coherence.

## 6 Limitation

This pilot study has several limitations that constrain how broadly the findings can be generalized. The overall dataset was modest (128 generated cases, 90 passing automated screening), and only a subset of the filtered pool received expert review (30 unique cases; 40 total ratings across 4 educators). These small samples—especially when broken down by model and condition—mean that observed differences between LLMs or conditions should be treated as suggestive rather than definitive, because a small number of atypical cases can disproportionately affect means, variance, and correlations. In addition, there was substantial imbalance in the number of cases produced and retained per model (e.g., GPT 4.1 contributing a large share of filtered cases, while Llama 4 Scout had a much lower pass rate), which reduces the fairness and statistical stability of cross-model comparisons.

Further limitations relate to the human and automated evaluation layers and the multimodal component. The expert panel was small and geographically bounded (MSK physiotherapy educators from Ontario, Canada), and educator judgments are context-dependent (e.g., documentation norms, imaging expectations, and thresholds for “usable” pedagogy vary by curriculum, region, and learner level). This context sensitivity likely contributed to lower agreement on criteria such as Medical Accuracy and Fidelity, where borderline plausibility decisions can reasonably differ across educators. The automated evaluation layer (DeepEval rubric and thresholds) was designed to screen for structure, alignment, and obvious safety concerns—not to guarantee clinical correctness—so weak or negative correlations between some automated metrics (notably Answer Relevance) and expert outcomes point to construct mismatch: “on-topic” and “instruction-following” are not equivalent to clinically coherent reasoning, allowing false-pass cases to reach experts.

## 7 Future Work

Future research should scale both generation and validation to strengthen generalizability and support condition- and model-specific conclusions by increasing the number of cases per condition, enforcing more balanced sampling across models, and expanding the expert panel in both size and diversity of teaching contexts (regions, program types, and learner levels). Methodologically, Phase 2 should evolve from primarily structural/topical screening into a clinical-coherence gate by adding explicit internal-consistency checks (e.g., mechanism-of-injury → symptom pattern, timeline and irritability coherence, appropriate handling of red flags and differentials, and imaging statements consistent with typical MSK pathways) and by redesigning rubrics toward reasoning-sensitive “process” evaluation rather than completeness/relevance of final JSON fields alone. Prompting should also be revised to reflect authentic MSK documentation—emphasizing pain behavior with movements/activities, distribution, irritability, and functional impact—because educators flagged overly specific and pedagogically unrealistic pain quantification (e.g., NPRS tied to a single muscle), and possibly implementation science frameworks to inform contextual and system level structure of the prompt strategy. Finally, the human evaluation layer should be made more rigorous by increasing double-rated overlap to stabilize IRR estimates, providing rater calibration materials, stratifying analyses by intended learner level to distinguish threshold differences from true disagreement, and adding a structured error taxonomy to make qualitative feedback more actionable for improving prompts and automated checks.

## 8 Conclusion

This study introduced PhysiCase, a pilot dataset and dual-layer validation workflow for generating synthetic MSK physiotherapy cases using frontier LLMs and evaluating them with both automated screening and expert educator review. Across 128 generated cases, the automated “LLM-as-judge” gate effectively filtered outputs for structural compliance, prompt adherence, and basic safety signals, yielding a smaller set of higher-potential cases for expert time. In human evaluation, educators rated the screened cases highly for usability, relevance, and realism, indicating that—once formatted, complete, and generally aligned—LLM-generated case narratives can meet many of the practical needs of MSK teaching and simulation.

At the same time, the findings clarify why expert review remains indispensable. Medical accuracy criteria showed the greatest variance and was the primary locus of expert-identified “nuance gaps,” particularly around mechanism coherence, imaging appropriateness, and unrealistic pain reporting that can subtly mis-teach clinical reasoning even when a case appears plausible. Cross-layer analyses further demonstrated that automated metrics such as answer relevance and prompt alignment are weak proxies for clinical correctness and pedagogical safety, whereas completeness is helpful but insufficient on its own.

Overall, the work supports a clear division of labor for scalable synthetic case development in MSK physiotherapy education. Automated evaluation can efficiently reduce the review burden by screening out structurally weak or misaligned outputs, but expert educators must remain the criterion standard for validating clinical coherence and teaching suitability. With targeted improvements to prompt design, reasoning-aware automated checks, more robust human validation, the PhysiCase approach can form the basis of a scalable, ethically defensible pipeline for building high-quality synthetic MSK case repositories for education and AI development.

## Data Availability

All data produced are available online at https://github.com/kwid-ai/PhysiCase

## Supporting information

### .1 Prompt for case generation

~~~
<Persona>
You are a seasoned physical therapist instructor at a reputable university. You are
↪ creating case studies for a specific topic.
</Persona>
<writing_style>
Write like a Senior professor with several years of industry experience
</writing_style>
<task>
You will be provided with a medical condition, using this condition, you
- A clinical history for a fictitious patient
- A title for the usecase are to create
- The muscle or group of muscles or bones affected by the condition.
- Numeric Pain Rating scale of the muscles out of 10 points – seperating primary and
↪ secondary pain complaint if applicable. The NPRS can be a range
- Aggravating factors if required
- Easing factors,
- 24 Hour Picture
- Past history if required
- Medications, if required
- Personal history, if required
Feel free to elaborate on the case study
</task>
<output_format>
Present the information in a JSON format following this structure
‘‘‘json
{
      “title”:”“,
      “clinical_history”:”“,
      “NPRS”:[
         {
             “context”:”P1”,
             “affected_area”:”“,
             “score”:”“
         }
      ],
      “aggrevating_factors”:”“,
      “Picture”:”“,
      “easing_factors”:”“,
      “past_history”:”“,
      “medical_history”:”“,
      “medications”:”“,
      “personal_history”:”“,
      “imaging”:”“
}
’’’
</output_format>
<example_output>
‘‘‘json
{
      “title”:”Elena with the case of hip persistent
      “clinical_history”:”Elena is a 56-year-old primary school teacher. 2/52 ago she was
      ↪ at work on a particularly difficult day with an aggressive student who fled the
      ↪ school the child was unaccounted for over an hour and there was a great deal of
      ↪ quick walking, running, stairs and squatting crouching associated with looking
      ↪ for the child. She noted the onset of left sided deep right groin pain over the
      ↪ course of the workday. She was particularly stiff and sore on rising after
      ↪ sitting. The pain worsened throughout the day, and she had a notable limp by the
      ↪ time she was walking to her car at the end of the school day. She had difficulty
      ↪ sleeping that night. The next AM her symptoms were worse on rising and she had
      ↪ difficulty getting upright and weight bearing on the affected side. She is
      ↪ currently experiencing (P1) deep groin and anteromedial thigh pain to the knee.
      ↪ In addition, there is pain over the lateral hip (P2). She has experienced
      ↪ intermittent clicking but has had no giving way. She lacks confidence on the
      ↪ limb after prolonged sitting or lying and will often shift and transfer weight
      ↪ rocking back and forth before ambulating independently.”,
      “NPRS”:[
         {
            “context”:”P1”,
            “affected_area”:”constant, deep groin and anteromedial thigh pain constant
            ↪ aching intermittent sharp/catching”,
            “score”:”4-6”
         },
         {
            “context”:”P2”,
            “affected_area”:”intermittent, lateral hip aching/burning”,
            “score”:”3”
         },
      ],
      “aggrevating_factors”:”walking (tolerance is no more than 30-45 minutes at a
      ↪ conservative pace and will be worse for a half hour following), standing 20-30
      ↪ minutes, sitting prolonged (noted on rising after 30-45 min), sitting cross
      ↪ legged for any period of time, descending stairs will aggravate P1 (immediately
      ↪ and will provoke sharp pain) and eventually P2 “,
      “Picture”:“[insert image of a person highlighting groin, anteromedial thigh and
      ↪ hip]”,
      “easing_factors”:“Aleve”,
      “past_history”:“Was involved in a head on collision lost control in bad weather and
      ↪ collided with a tree 14 months ago. Hip was sore for about 4/52 and then settled.
      ↪ Has experienced intermittent short episodes of groin tightness, buttock cramping
      ↪ and soreness after being in a squat or standing for a prolonged period. Symptoms
      ↪ have never limited function in the past.”,
      “medical_history”:“hysterectomy 3 years ago secondary to fibroids but generally well.
      ↪ Has gained approximately 30 pounds over the last few years following surgery.
      ↪ Not really an exercise person but will walk and enjoys gardening.”,
      “medications”:“Aleve prn”,
      “personal_history”:“married with three children. Two children are married and live
      ↪ in nearby cities. Youngest son suffers from schizophrenia and lives with the
      ↪ patients home, as they are unable to manage in an independent living environment.
      ↪ Their child was diagnosed three years ago following an escalation of symptoms for
      ↪ two years. This has been a significant issue within the family and resulted in a
      ↪ significant strain on many of the family relationships as they support their son
      ↪ through this time.”,
      “imaging”:“none to date”
}
’’’
</example_output>
~~~

### .2 Prompt for DeepEval - Completeness metrics

~~~
  metric = GEval(name=“overall_performance”,
               criteria=“Determine whether the actual output is factually correct,
               ↪ complete and comprehensive”,
               evaluation_steps=[
                     “Check if the output contains all required fields as per the
                     ↪ output format.”,
                     “Verify that the content is relevant to the provided medical
                     ↪ condition.”,
                     “Assess the clarity and comprehensiveness of the response”,
                     “Provide reasons for the score, emphasizing the importance of
                     ↪ clinical accuracy and patient safety.”
               ],
     evaluation_params=[LLMTestCaseParams.ACTUAL_OUTPUT], rubric=[
        Rubric(score_range=(0,2), expected_outcome=“Factually incorrect.”),
        Rubric(score_range=(3,6), expected_outcome=“Mostly correct.”),
        Rubric(score_range=(7,9), expected_outcome=“Correct but missing minor
        ↪ details.”),
        Rubric(score_range=(10,10), expected_outcome=“100% correct.”)]
     )
~~~

## A Expert review comments and taxonomy categorization

### A.1 Category 1: Reductive Logic

**Table 14.**
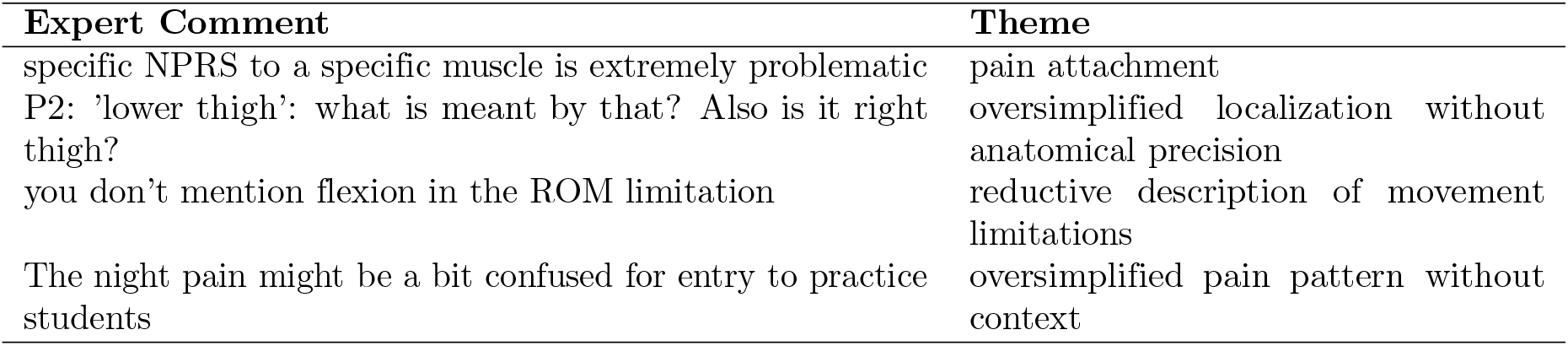
Category 1.

### A.2 Category 2: Biomechanical Inconsistency

**Table 15.** Category 2.

| Expert Comment | Theme |
| --- | --- |
| Mechanism is problematic |  |
| Using a cane seems out of place | aid doesn't match functional presentation |
| He seems in great shape otherwise. Using a cane seems out of place |  |
| Should give info on walking aid (is he using crutches) | inconsistency in mobility aid description |
| Lacks context: did he stopped working since the injury? WSIB? Are there cuts? | timeline/functional status inconsistency |
| Clinical history: should add more context. The injury could have arrived at the end of the last day of the tournament | mechanism timing mismatch |
| Easing factors: keeping the finger elevated. Need more info, how? Above the heart? | biomechanically vague positioning |
| I'm not sure how realistic it would be for a college level athlete to wait 2 days to see someone | behavioral inconsistency with population |
| Usually due to an acute change in activity... so I wonder if the weight gain as a provocative factor isn't fully realistic | mechanism inconsistent with typical presentation |

### A.3 Category 3: Administrative/Contextual Gaps

**Table 16.** Category 3.

| Expert Comment | Theme |
| --- | --- |
| Need information on ambulation (with what type of aid? Can he walk only with the prosthesis? | missing functional context |
| Personal history: concerns about finance but this is a work injury so he should be on WSIB | insurance/context inconsistency |
| We can infer he is over weight, but there is no picture or objective indicator ie BMI | missing objective data |
| Also should could report functional problem with him restriction ie. putting on socks | missing functional limitation documentation |

## Acknowledgments

This work was made possible through the support of Label Studio Academic Program.

